# Lung functions among children and adolescents with sickle cell disease receiving care at Jaramogi Oginga Odinga Teaching and Referral Hospital Kisumu, Kenya

**DOI:** 10.1101/2024.08.22.24312427

**Authors:** Fredrick O. Olewe, Constance N. Tenge, Irene K. Marete

## Abstract

Sickle cell disease is a genetic disorder associated with lifelong symptoms of anemia, vaso-oclusive crisis and organ damage. Pulmonary complications and declining lung functions are contributors to morbidity and mortality. Determination of lung functions can enable early diagnosis and institution of appropriate intervention.

The objectives were; To determine the prevalence, patterns, and the factors associated with abnormal lung functions in children and adolescents aged 6-17 years with confirmed diagnosis Sickle Cell Disease.

This was a cross sectional study done at Jaramogi Oginga Odinga Teaching and Referral Hospital in Kisumu, Kenya. A total of 138 participants were recruited. A structured data collection tool was used to collect Socio-demographics and clinical characteristics. The spirometry was done using NDD Easy-On PC spirometer and results entered into database, cleaned and analyzed for proportions, frequency, mean, range and odds ratio at 95% confidence level.

Females were 65(47%), males 73(53%), and the mean age was 10.99 years with SD of 3.15. A total of 79(57%) were from rural and fuel used was fired wood 41%. Hospital admission were 84(61%), vaso-oclusive crisis 104 (75%), acute chest syndrome 61(44%) and blood transfusions 56(40%) in the last 12 months. Prevalence of abnormal lung functions was 28 % (39), restrictive pattern 26(67%) and obstructive pattern 13(33%). Urban setting (OR 24.101 p value 0.163), being female (OR 18.911 p value 0.069), and blood transfusion (OR 11.683 p value 0.195) had high odds ratio while, stable hydroxyurea use (OR 0.525 p value 0.678) and hospital admission (OR 0.048 p value 0.121) had low odds ratio, but not statistically significant.

One third had abnormal lung function mostly restrictive followed by obstructive pattern. Residence in urban setting, being female and blood transfusion had high odds ratio, while stable hydroxyurea use and hospital admission had low odds ratio but no statistically significant.

## Introduction

Sickle cell Disease (SCD) is a genetic disorder of the hemoglobin molecule that forms part of the red blood cell. The red blood cell is made up of a hem group and four globin chains which comprise of two beta and two alfa chains (B2a2). The gene clusters on chromosome 16 and 11 control globin chain production. The allele which is responsible for production of SCD is autosomal recessive and is found on the short arm of chromosome 11 (1).

In Sub Saharan Africa, around 240,000 children are born with the disease each year (2). Most of these children die without ever being diagnosed as there is limited newborn screening and early treatment in Sub-Saharan Africa. SCD is responsible for 50-90% of under-five mortalities in low income countries. This is because there is limited data on the natural history of the disease in our setting which makes early identification and treatment of the complications difficult leading to poor prognosis (3)

It is predicted that the numbers of children with this disorder will steadily increase over the next 40 years if mechanisms are not put in place to control the disease. Given this predicted rise in numbers of children with SCD, major investment in clinical research is necessary to fully understand the natural history of sickle cell disease in our setting and the associated respiratory complications (4).

Children with homozygous state (Hb SS which usually have a more severe course of the disease or Hb SC with a milder course of the disease) are usually symptomatic with lifelong symptoms. which can be acute or chronic with chronic organ damage which can present as kidney failure, stroke, stunting cardiomyopathy and chronic lung disease which present with respiratory symptoms (5) (3)

Usually, the severity of the disease is attributed to a number of factors such as genetic modifiers, nutritional status, access healthcare among others (6). However, the real burden and the prevalence of associated morbidities such as obstructive and restrictive lung disease which can eventually progress to chronic lung disease remains elusive (4).

There have also been contradicting findings from previous studies on whether the pattern of lung function abnormality contributes to future morbidity and mortality among SCD patients. In contrast, it has been argued that, the pattern of abnormal lung function test is not associated with previous or future painful crisis or acute chest syndrome and that, the abnormal lung functions are sometimes over estimated in certain circumstances (7).

It has been difficult comparing results between older studies which is hindered by the fact that there were many different reference values and criteria used to define lung function abnormalities (8)

Acute and chronic lung complications in sickle cell disease are major causes of morbidity and mortality, accounting for up to 20-30% of all sickle cell deaths. This indicates that screening should be done early and required interventions instituted in order to reduce the burden of respiratory related complications among sickle cell patients (9)

The most prevalent abnormal spirometry findings among children and adolescents with SCD varies globally. In the US in a study done by Cohen and colleagues, noted that 30% of children had an abnormal lung function pattern. The most common abnormal pattern was obstruction which was found in 16%, with fewer patients having restriction at 7% (7). In Italy, Tassel found out that the most prevalent pattern was restrictive at 13% with obstruction at 5%, which is similar to what has been noted in some of the African countries (10)

In Sub Saharan Africa, which is a low income setting, restrictive spirometry pattern is the most prevalent pathological pattern. However, among pediatric patients with SCD living in high-income countries, obstructive lung function is more frequent than restrictive. This is a study which was done to compare the abnormal lung functions among children with SCD in Africa and United Kingdom (Arigliani, et al., 2019)

In the general populations, it has been established that a number of factors affect lung function, both environmental and intrinsic factors. In addition, it has been noted that lung function decline over time among children living with SCD. Regarding risk factors for abnormal lung function in children with SCD, it has been pointed out that there is an association with history of acute chest syndrome (ACS), whereas the rate of painful crises seemed to be unrelated to respiratory abnormalities, it has also been observed that patients with more than 3 episodes of ACS had lower FEV1 than the predicted value (12), (13), (14).

In addition, Malnutrition was also associated with increased risk of restrictive spirometry pattern in children with SCD from Central Africa, while high body mass index (BMI) percentile was found to be an independent predictor of increase in forced expiratory volume at one second (FEV1) in patients from High Income Countries. Furthermore, Sex as an independent variable has been noted to have no significant influence on the respiratory function abnormalities in children with sickle cell disease in the same study. Arigliani and colleagues also noted that lung function decline with age in both children from low income and high income countries with differences in the patterns of abnormities (12) (8)(15)(16), (17)

In a study done in West Africa, ACS, hydroxyurea use and painful crisis were found to be significantly associated with changes in lung functions. Furthermore, the use of hydroxyurea as a disease modifying drug in sickle cell disease reduces the progression of loss of lung function in children with SCD (18) (19). Additionally, a History of asthma and wheezing has also been associated with obstructive lung function abnormality as was observed in a study by Manuel and colleagues (20), (21).

## Methods and materials

### Study site

This was a descriptive cross-sectional hospital-based study carried out at Jaramogi Oginga Odinga Teaching and Referral Hospital (JOOTRH) in Kisumu a level five hospital in western Kenya. Recruitment was done at the two outpatient clinics. The clinic at the Obama Children’s hospital catered for children aged 6 to 13 years while those who are between 14 to 17 years were recruited from the hemato-oncology clinic within the same hospital which has a well-established sickle cell clinic.

### Study population

Children with SCD receiving care at JOOTRH

### Study period

The study was done over a period of six months from 1^st^ October 2023 to 30^th^ March 2024

### Eligibility criteria

#### Inclusion criteria

1. Children between 6-17 years with clinical features of SCD and with a confirmatory test and receiving care at JOOTRH whose parent or guardian consented and those who above 7 years gave a written assent

#### Exclusion criteria

1. Confirmed cases of or on treatment for pneumonia, pulmonary tuberculosis or any other respiratory disease at the time of recruitment
2. Children who were not able to follow instruction for the study procedures due to illness or any other reason
3. Children who were on bronchodilator or steroids for active treatment of asthma

#### Sample size

The sample size was calculated using the Fisher’s formula based on a p of 0.3 from the study by Cohen which showed a prevalence of abnormal lung function at 30% (7).

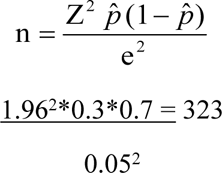

Given that the total number of unique children above 6-17 years seen per month was approximately 30, therefore, the approximate total population of interest during data collection period was estimated to be 240 children.

Corrected sample size for finite population

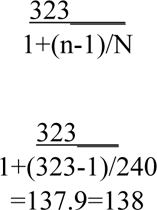

### Recruitment

The Principal Investigator (PI) and the research assistants were trained on spirometry by a Respiratory Society of Kenya (RESOK) trainer and certified to perform and interpreted the spirometry results. After being attended to by the clinician on duty for routine clinic visit, potential participants were directed to the PI or the research assistant for screening who identified children who met the study requirements. Parents or guardians who were willing to let their children join the study proceeded to give written consent and written assent was obtained from children above 7 years. The PI or the research assistant took a short medical history of the participants including demographics such as age and sex, residence rural or urban, cooking space whether indoor or outdoor, fuel used for cooking whether firewood, gas or charcoal. Medical information was also obtained which included hydroxyurea use, number of vaso-occlusive crisis (VOC), ACS, blood transfusion, and hospital admission in the past 12 months. Anthropometric measurements such as weight and height was obtained and used to calculate BMI and recorded on a structured data collection tool which was validated before the study. Where information was not clear, this was clarified from the patients’ folders.

Two NDD Easy-on PC spirometer device were available for the study where one acted as a backup. The spirometry test was presented as a blowing game with computer incentives of blowing off the candles on a birthday cake first for purposes of practice and during the actual procedure where blowing off all the candles corresponded with achieving the required forced expiration. The participants were encouraged to blow and where necessary demonstration was done alongside the participant.

The patients were put in a sitting position with their arms resting on the arm rest of the chair and their back straight and given time to relax and be composed before the spirometry procedure. A clean mouth piece was attached to the spirometer device, patient’s nose clipped and he or she was instructed to take in normal three breathes before taking in a deep breath and then blowing out forcefully for 6 seconds, as the instructor verbally encouraged the patient and the patient looking at the candle on the screen. The procedure was repeated 3 times to obtain the highest values with minimal curve variability, which was in conformity with standard international guidelines of European Respiratory Society and American Thoracic Society in terms of quality and reproducibility (23). Quality control was done to ensure consistency and the best values were used for interpretation of the results in comparison to the predicted reference values. Where there was difficulty interpreting the results, a pulmonologist was consulted for assistance

### Independent variables

Some of the independent variables in the study were age, sex, BMI which was categorized as; normal, underweight, overweight or obesity, vaso-occlusive crisis, acute chest syndrome, blood transfusions, number of hospital admissions, stable hydroxyurea use, history of asthma or wheeze, type of cooking fuel used, residence either rural or urban and cooking area either outdoor or indoor.

### Dependent variable

The dependent variable was abnormal lung function of which three types of outcomes were considered which were; restrictive i.e. lower FEV1<80% of lower limit of normal and obstructive i.e. lower FVC <80% or FEV1/FVC ratio <70% of the lower limit of normal or mixed with binary out comes for each and was coded as 0 absent or 1 present for each case.

### Data Management and Analysis

#### Data entry and cleaning

Data was entered into database and cleaned to confirm accuracy and conformity with the source document before analysis.

#### Data analysis

Data analysis was done according to the statistical analysis plan (SAP) in place to compute the frequency and proportion for categorical data, mean, SD and range for continuous data. A bivariate analysis was done for association, odds ratio at 95% confidence level. Fishers exact tests where applicable to test for statistically significance.

#### Ethical Approval

The study procedures were started after obtaining ethical approval from the Institutional Research Ethics Committee (IREC) of Moi University/ Moi Teaching and Referral Hospital and from Jaramogi Oginga Odinga Teaching and Referral Hospital Institutional Scientific Ethics and Research Committee (ISERC). Administrative approval was obtained from the Hospital allowing the conduct of the study within the facility.

Written consent was obtained from parents/guardian of all the children below 18 years and assent from children above 7 years.

#### Confidentiality

The participant’s information was de-identified to remove any personally identifiable information in order to maintain confidentiality. The folders were kept in lockable drawers which were accessible to the PI and other authorized study staff only

## Results

A total of 151 participants were screened, 13 of them were screen excluded while 138 met the inclusion criteria and considered for the final analysis

### Socio demographic characteristics

#### Characteristics of the participants’ care giver

Out of 138 participants, 103(75%) of the participants were brought to the hospital by their mothers, 13 (9%) were brought by their fathers while only 8(6%)were brought by others who were neither their mother, father or guardian. Out of the total number of parents, guardians and others who brought the participants to the hospital, 59(43%) of the parents or guardians of the children surveyed had attained secondary education, 42(30%) had Tertiary level while 3(2%) had no education.

#### Socio-demographic characteristic of the participants

Table 1 shows the demographic characteristics; Sex distribution shows males were of 73(53%) while the female were 65(47%).

**Table 1:**
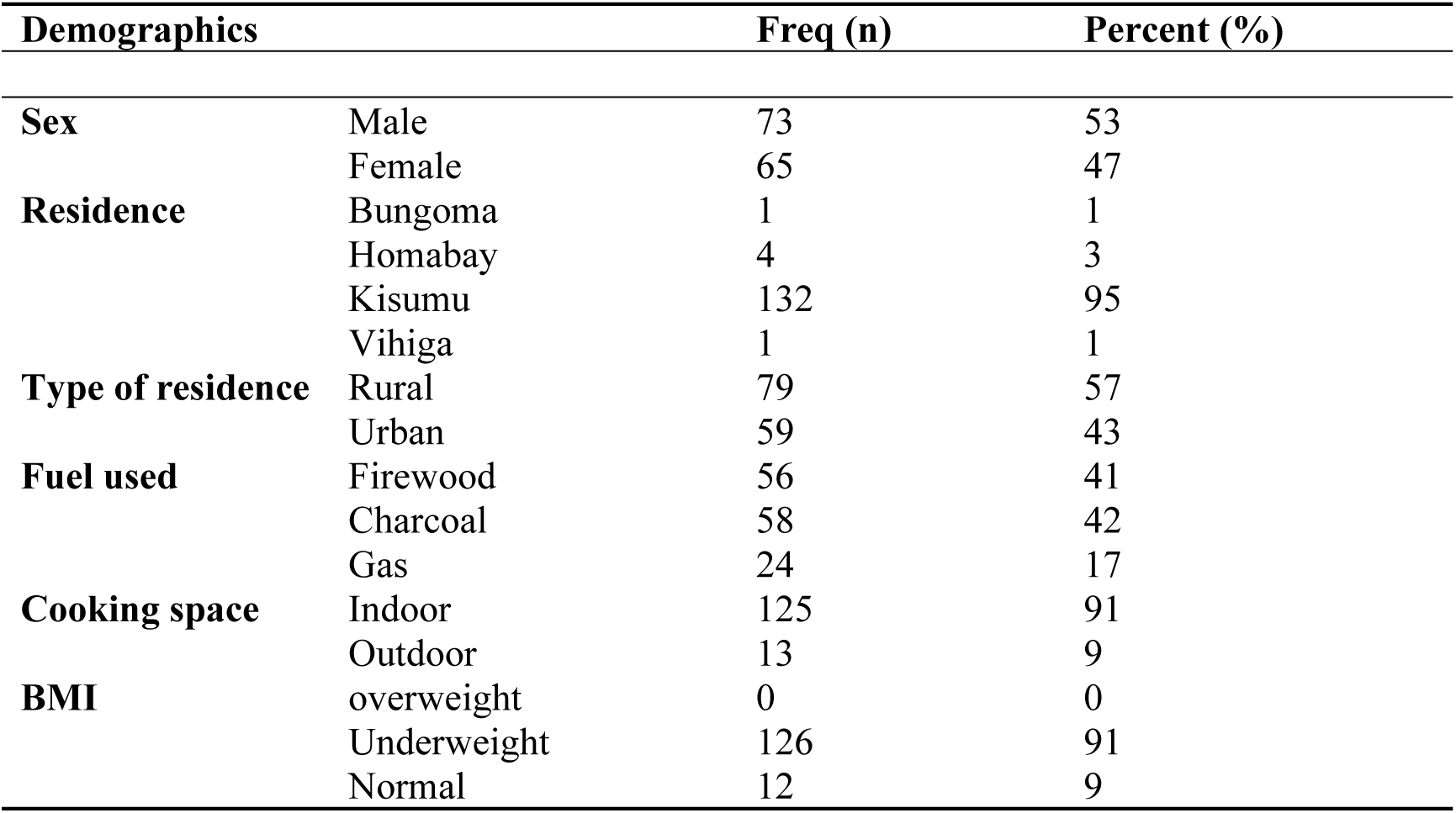
sociodemographic characteristics.

#### Anthropometric measurements

Anthropometric measurements showed that the mean age was 10.99 years with an SD of 3.15. The mean weight of the children was 29.06 kg (with the minimum weight of 14.1 kg and maximum weight of 62 kg) and SD of 9.8. The children had a mean height of 138.19 cm, (minimum height was108 cm and maximum height as 178 cm) and SD of 15.5.

#### Clinical characteristics

Looking at the clinical features in table 2, 102(74%) of the children reported being on stable dose of hydroxyurea within the las 3 months, 34(25%) reported no VOC while 71(51%) had 1-2 VOC, additionally,77(56) had no ACS while 52 (38%) had one to two ACS in the past 12 months. In addition, 82(59) had received no blood transfusion and only 10(4%) had received more than three blood transfusions in the past 12 months, while 84(61%) had been admitted to the hospital within the last 12 months

**Table 2.** patients clinical factors.

| Clinical characteristics | Freq (n) | Percentage |
| --- | --- | --- |
| **Stable hydroxyurea dose** |  |  |
| **Yes** | 102 | 74 |
| **No** | 36 | 26 |
| **No of VOCs within 12 months** |  |  |
| **None** | 34 | 25 |
| **one-Two** | 71 | 51 |
| **>three** | 33 | 24 |
| **No of ACS within 12 months** |  |  |
| **None** | 77 | 56 |
| **one-Two** | 52 | 38 |
| **>three** | 9 | 6 |
| **No of Blood transfusion within 12 months** |  |  |
| **None** | 82 | 59 |
| **One-two** | 46 | 33 |
| **>three** | 10 | 4 |
| **Admission history within 12 months** |  |  |
| **Yes** | 84 | 61 |
| **No** | 54 | 39 |

#### Prevalence of abnormal lung function

Out of the total number of the children, 39(28%) of them had abnormal lung functions, while 99(72%) had normal lung function test

#### Patterns of abnormal lung functions

Table 3 shows patterns of abnormal lung function. Out of the 39 children with abnormal lung functions 26(67%) had restrictive lungs function pattern while the rest 13(33%) had obstructive lung function pattern with none of the children having obstructive pattern.

**Table 3:** patterns of abnormal lung functions.

| **Pattern of abnormal lung function** | **n** | **Percentage (%)** |
| --- | --- | --- |
| **Restrictive** | 26 | 67 |
| **Obstructive** | 13 | 33 |
| **Mixed** | 0 | 0 |

#### Factors associated with abnormal lung functions

Table 4 shows factors associated with abnormal lung functions. The findings showed that being female appears to have a substantially high odds ratio of 18.91 compared to males, though the p-value 0.069, 95% CI (0.794, 450.23). There is no significant association between weight status (underweight or normal) with abnormal lung functions, normal weight low odds ratio of 0.3036 and a p-value of 0.301 95% CI (0.032, 2.902). Living in an urban setup had remarkably high odds ratio 24.10 compared to rural settings, with a p-value 0.163 95% CI (0.276, 2104.596).

**Table 4:** Factors associated with abnormal lung functions.

|  |  | Odds Ratio | P Value | [95% Conf. Interval]<br>Lower, upper |
| --- | --- | --- | --- | --- |
| **Sex** | Male | REF |  |  |
|  | Female | 18.911 | 0.069 | 0.794 , 450.25 |
| **Weight** | Underweight | REF |  |  |
|  | Normal | 0.304 | 0.301 | 0.032, 2.902 |
|  | overweight | 0 |  |  |
| **Type of residence** | Rural | REF |  |  |
|  | Urban | 24.102 | 0.163 | 0.276, 2104.596 |
| **Type of fuel used at home** | firewood | REF |  |  |
|  | charcoal | 0.196 | 0.389 |  |
|  | Gas | 0.134 | 0.486 |  |
| **No. of ACS in past 12 months** | 0 | REF |  |  |
|  | 1-2 | 2.444 | 0.533 | 0.147, 40.578 |
| | $\geq 3$ | 1 (empty) | | |
| **No of Blood Transfusions received in the last 12** | 0 | REF |  |  |
|  | 1-2 | 11.683 | 0.195 | 0.285, 479.720 |
| | $\geq 3$ | 1 (empty) | | |
| **Stable hydroxyurea dose for the last 3 months** | No | REF |  |  |
|  | Yes | 0.525 | 0.678 | 0.025, 10.927 |
| **admission in the past in the last 12 months** | No | REF |  |  |
|  | Yes | 0.048 | 0.121 | 0.001, 2.225 |

#### Clinical factors and patterns of abnormal lung functions

Table 5 shows clinical factors and patterns of abnormal lung functions where more than 3 blood transfusion in 12 mothers showed statistically significant association with both restrictive and obstructive abnormal patterns of lung function.

**Table 5:** Clinical factors and patterns of abnormal lung functions.

| Factors |  | Restrictive<br>(Low FVC) | Obstructive<br>(Low FEV1) | P-value |
| --- | --- | --- | --- | --- |
| **No of Vaso-Occlusive Crisis** | 0 | n(%) | n(%) | 0.614 |
|  | 1-2 | 7(70) | 3(30) |  |
|  | 3 | 13(85) | 2(15) |  |
| **Number of Acute chest syndrome (ACS)** | 0 | 11(75) | 3(25) | 0.816 |
|  | 1-2 | 16(78) | 4(22) |  |
|  | 3 | 12(73) | 5(27) |  |
| **Number of blood transfusions received** | 0 | 2(100) | 0(0) | **0.031** |
|  | 1-2 | 14(80) | 3(20) |  |
|  | 3 | 16(82) | 4(20) |  |
| **Patient on stable hydroxyurea dose for the last 3 months** | Yes | 0(0) | 2(100) | 0.566 |
|  | No | 22(79) | 6(21) |  |
| **History of asthma** | Yes | 8(73) | 3(27) | 0.573 |
|  | No | 1(100) | 0(0) |  |
| **History of Wheezing** | Yes | 29(76) | 9(24) | 0.314 |
|  | No | 3(100) | 0(0) |  |
| **Admitted in the past 12 month** | Yes | 27(75) | 9(25) | 0.434 |
|  | No | 19(79) | 5(21) |  |

## Discussion

### Prevalence of abnormal lung functions

Children living in low and middle income countries still have a substantial burden of abnormal lung functions. This was demonstrated in this study where 39(28%) of the children had an abnormal lung functions mostly of restrictive followed by obstructive pattern. Factors such us being female, blood transfusion and living in urban setting had high odds ratios while hospital admissions and stable hydroxyurea use had very low odds ratios.

There is poor access to health services and noncompliance to the sickle cell treatment leading to complications such as VOC, ACS and hypoxic injuries. These frequent insults from hypoxia VOC and ACS contribute to the interstitial lung damage leading to abnormal lung functions (8). There was also a female predominance where out of 66, 20(30.3%) had abnormal lung function. This female predominance can be explained by the cultural practices of cooking with firewood in an enclosed cooking space as women are expected to cook hence exposure smoke which leads to lung injuries hence abnormal lung functions. The findings are similar with the study done in the USA by Cohen et al that found a prevalence of 30% among children and adolescents (7). However, in a study done by Dei-Adomakoh and colleagues in adults with sickle cell, there was 70.4% prevalence which indicated that the decline in lung function among children is a continuous process and worsens as they grow older (14). The difference can be explained by the fact that the study populations were different in characteristic majorly adults as oppose to children and adolescents in our case.

In Nigeria, in a study that was done by Arigliani to compare lung functions among Nigerian children and those from the United Kingdom, there was a prevalence of 38% in Nigeria, and 27 % in United Kingdom. This is slightly higher in Nigeria than our study which was at 28%, but finding in the UK were similar to this study despite the fact that this was a cohort study where the participants were followed up and UK has a better health care system than Africa which alludes to the fact that developing abnormal lung function could be intrinsic to having sickle cell disease. In this Study also, confounders were excluded as appropriate (11). These findings cannot over emphasize the need for including lung function test as part of routine care for children and adolescents with sickle cell disease

In Kuwait, A Adekite found a similar prevalence of 28.5%. Besides, they used a different phenotype of HbSC with very high fetal hemoglobin levels with mild symptoms and still had comparable prevalence to this study which involved HbSS phenotype. This equally showed that there is more work to be done to establish the exact mechanism of lung damage in sickle cell disease (24)

### Patterns of abnormal lung function in children with SCD

There has been a contentious argument as to what is the most prevalent pattern of abnormal lung function among children with sickle cell disease and as to whether this varies with regions. In this study, we found out that restrictive pattern was the most prevalent and double at 26(67%) while obstructive pattern was 13(33%). There was no mixed pattern observed in this study. The restrictive pattern seen in this study can be explained by the fact that most of the participants were underweight which is associated with restrictive lung functions. This is in contrast to a study by Cohen, where they found out that the most prevalent pattern was obstructive at 16% (7). This was possibly due to the urban setting of their studies as previously it has been shown that children in such set up are more predisposed to hyperactive airway disease. In Ghana, Kuti and Adegoke found that most of the children had restrictive lung disease followed by obstructive and a few had mixed pattern (25). This is in congruent to this study where we found out a similar trend except for mixed pattern which was not found among these children, which could be because we used only spirometry which was not adequate in characterizing the patterns further.

There seem to be regional variation of the patterns as in the study done by Manuel et al which found exact reverse of our findings in this current study. In their study, the most prevalent pattern was obstructive at 19% followed by restrictive at 9% (20), they equally did not report on the mixed pattern. Their study setting was more urban which is similar to the study by Cohen and colleagues but with different results. In this study, reversibility testing was not done to further characterize the obstructive pattern into either reversible or irreversible which would give a glimpse of the possible cause as this was outside the study aims.

The previously differing opinions of the pattern of abnormal lung functions among children with sickle cell disease is slowly taking shape as more studies indicate that in Africa and some European countries, the most prevalent pattern is restrictive lung disease. This pattern can be explained by the repeated insults to the lungs as a target organ as had been alluded to earlier. Furthermore, it was noted by Kuti and Adegoke in Nigeria in their study that 22.1% of the children with SCD had restrictive pattern while 5.8% had obstructive and only 1.9% had mixed pattern (25). Though the findings are slightly different, it gives a similar picture to the findings in this study where 19% had restrictive, 9% obstructive with none having mixed pattern.

### Factors associated with abnormal lung functions in children with SCD

It is now known that lung functions decline over time among children living with Sickle Cell Disease, however, there is paucity of data on some of the contributing factors to this rapid decline. However, the factors that contribute to this decline still remains elusive to the medical fraternity. Different studies have found different factors to be contributory, furthermore, most factors are usually not statistically significant despite the high odds ratio. This was no different from this study.

In this study being female had an increased risk of having an abnormal lung function compared to male OR 18.91 CI 0.7943,450.25, however, this was not statistically significant with a p value of 0.069. This is similar to the study done in Central Africa by Arigliani which associated being female with increased risk of having abnormal lung functions (12). The increased likelihood in females compared to males in this study can be explained by the fact that in greater Africa and mostly in rural areas cooking is done indoors with firewood which produces a lot of smoke by females increasing their exposure to pollutants leading to lung damage hence developing the abnormal lung functions. This was in contrast to the previous studies which showed that sex has no influence on the risk of having an abnormal lung function among children with sickle cell disease (9), (26). This difference could not be explained since there were more males in this study compared to females at 73(52.55%) and 65(47.45%) respectively. However, it can be hypothesized that the difference could be attributed to the cooking environment where mostly occupied by females.

When it comes to the BMI, most studies have found an association between obesity and abnormal lung functions mostly obstructive lung disease as in the case of a study by Koumbourlis where BMI had a statistically significant influence with a p value of 0.02 (27). In this study having a normal BMI was shown to be protective, however, this was not statistically significant with a p value of 0.301, with OR of 0.304 at 95% CI (0.0318, 2.9019). This similarity is due to the fact that we used similar populations of study. In another study however, BMI was found to have no association with abnormal lung function (14). This difference could be explained by the fact that they used slightly different study population.

In this study, it was postulated that urban residence may have some significant contributions in children with sickle cell anemia, this could be similar to populations without SCD. This was attributed to the industrial and motor vehicle emissions causing air pollution in town centers which contributes to lung injury. It was found out that living in urban set up had an increased risk of having an abnormal lung functions at more than 24 times compared to rural set up. However, this was not statistically significant OR 24.1015 CI 95% (0.2760, 2104) with a p value of 0.163.

In Nigeria, use of solid fuels was found to have no significant association with abnormal lung functions in children with sickle cell disease (28). This was similar to this study where it was found out that type of fuel had no significant association with abnormal lung functions p value 0.389 for charcoal and 0.486 for gas. However, compared to use of fired wood, charcoal and gas were protective with OR of 0.1959 and 0.1344 respectively. This can be attributed to the amount of smoke produced by firewood within an enclosed indoor cooking place which predisposes the house occupants to air pollutions hence increasing chances of having lung damage and abnormal lung functions.

Given the hypothesized mechanism of lung injury leading to fibrosis and interstitial lung injury with ultimate impaired lung function in children with SCD, the number of Acute Chest Syndrome (ACS) has been associated with the decline in lung function in this kind of population (29). In this study, children who had more than 2 episodes of ACS in the past 12 months had a slightly increased likelihood of having an abnormal lung functions twice as high compared to those who had none, however, this was not statistically significant OR 2 .44 CI (0.1473, 40.5778) at 95% confidence level. In a study by Arigliani, having more than 3 episodes of ACS was associated with impaired lung function (12). However, Cohen and colleagues found no association of ACS with abnormal lung functions. This difference can be explained by the fact that Cohen study was done in a high income country with good health care services to children with SCD which limited the effect of ACS due to timely and adequate management of the acute complications of patients with sickle cell disease. Kuti also found out that ACS was associated with abnormal lung functions in Nigerian children with SCD with an OR of 8.5 (25), (7).

Sometimes children with SCD, can have complications which warrant urgent transfusions. This can be occasional or at times they can be dependent on chronic transfusion. As an independent factor, transfusion was associated with abnormal lung functions with a p value of 0.195. However, in the presence of other factors, children who received more than 2 transfusions within the last 12 months had greatly increased risk of having an abnormal lung function even though it was not statistically significant OR 11.683 95% CI (0.285, 479) P value 0.195 this increased likelihood may not be due to transfusion itself but could be attributed to other issues such as noncompliance to medications and clinic appointments predisposing them to multiple complications which contribute to their abnormal lung functions.. This was different from the study by Adekile in Nigeria which found out that that there was no association between FEV1 and transfusions (24). This difference was because they narrowed only to the effect on FEV as opposed to generally looking at the abnormal lung function as was the case in this study.

The mainstay of therapeutic management of SCD patient is by use of hydroxyurea which is based on the weight and patients’ response. Lung functions have been shown to improve after starting hydroxyurea treatment (18). This raised the questions as to whether hydroxyurea use can slow or prevent the development of abnormal lung function. In this study, being on a stable dose of hydroxyurea for three months shown to be protective, however, it was not statistically significant with an OR 0.566 at 95% CI (0.02, 10.92) with a p value of 0.678. This was similar to a study by J. Stewart who found no significant association with a p value 0.24 (30). Similarly Arigliani found no significant association in Central Africa (12). This shows that there is still work to be done in order to ascertain the contribution of hydroxyurea to the decline in lung functions among children with SCD. In a meta-analysis by Taksande, hydroxyurea use was found to be protective, but not statistically significant which is similar to this study (21).

Following the devastating complications of SCD, most children needs frequent and repeated hospital admission for appropriate and proper care. This is not possible for some families from low income countries thus leads to inadequate medical care. In this study, hospital admission within the past year was protective but not significant OR 0.047 P value 0.121 CI (0.001, 2.224). this can be explained by the fact that children with more hospital admissions received adequate and appropriate medical care which reduced their chances of developing the complications associated with development of abnormal lung functions. This was different from a study by Kuti which found a significant association of abnormal lung functions with hospitalization p value <0.05 in Nigeria (25), but similar to the findings of the study by Arigliani and colleagues (8). It is therefore necessary for improved health care services for children and adolescents with SCD in order to improve their general well-being and reduce risk of developing an abnormal lung functions. This study had a limitation of wider confidence interval which can be improved in future by doing cohort studies in Africa with larger sample sizes.

## Conclusion

The prevalence of abnormal lung function among children with Sickle Cell Disease was 28%

The most prevalent pattern of abnormal lung function was restrictive followed by obstructive and no mixed pattern

Urban setting, being female and the number of blood transfusion received within the past 12 months were associated with high odds ratio while hospital admissions and stable hydroxyurea used were associated with low odds ratio.

## Data Availability

Data will be deposited in a public depository, currently can be obtained by request from the corresponding author

## Acknowledgment

I would like to thank Prof. Bernhards Ogutu, Dr. Vincent Were, Dr. John Orimbo, Dr. Allan Otieno, Caroline Atieno, Pamela Atieno Nashon Okello, Sophy Omondi, Grace Omukoko, Ishmael Sports and Jane Olum for their valuable contribution towards the realization of the study goals.

## Notes

### Competing Interest Statement

The authors have declared no competing interest.

### Clinical Trial

NA

### Funding Statement

No funding was received for this work

### Author Declarations

Institutional Review Ethics Moi University

